# Using group norms to promote acceptance of HIV testing during household tuberculosis contact investigation: A household-randomized trial

**DOI:** 10.1101/2024.05.02.24306703

**Authors:** Mari Armstrong-Hough, Amanda J. Gupta, Joseph Ggita, Joan Nangendo, Achilles Katamba, J. Lucian Davis

**Affiliations:** Department of Epidemiology, New York University School of Global Public Health, New York, NY, USA; Department of Social & Behavioral Sciences, New York University School of Global Public Health, New York, NY, USA; Uganda Tuberculosis Implementation Research Consortium, Walimu, Kampala, Uganda; Epidemiology of Microbial Diseases, Yale School of Public Health, New Haven, CT, USA; Johns Hopkins Bloomberg School of Public Health, Baltimore, MD, United States; Clinical Epidemiology Unit, Department of Medicine, Makerere College of Health Sciences, Kampala, Uganda; Pulmonary, Critical Care, and Sleep Medicine Section, Yale School of Medicine, New Haven, CT, USA

**Keywords:** Home-based HIV testing, TB Contact investigation, Misperceived norms, Stigma

## Abstract

**Background:** HIV status awareness and linkage to care are critical for ending the HIV epidemic and preventing tuberculosis (TB). Among household contacts of persons with TB, HIV greatly increases the risk of incident TB and death. However, almost half of household contacts in routine settings decline HIV test offers during routine contact investigation. We evaluated a brief social-behavioral norming intervention to increase acceptance of HIV testing during household TB contact investigation.

**Methods:** We carried out a household-randomized, controlled trial to evaluate the effect of the norming strategy among household contacts of persons with pulmonary TB in Kampala, Uganda (ClinicalTrials.gov #NCT05124665). Community health workers (CHW) visited homes of persons with TB to screen contacts for TB symptoms and offer free, optional, oral HIV testing. Households were randomized (1:1) to usual care or the norming strategy. Contacts were eligible if they were ≥ 15 years old, self-reported to be HIV-negative, and living in a multi-contact household. The primary outcome, the proportion of contacts accepting HIV testing, was analyzed using an intention-to-treat approach, using a mixed-effects model to account for clustering by household. We assessed HIV testing yield as a proportion of all contacts tested.

**Results:** We randomized 328 contacts in 99 index households to the norming strategy, of whom 285 (87%) contacts were eligible. We randomized 224 contacts in 86 index households to the usual strategy, of whom 187 (84%) contacts were eligible. Acceptance of HIV testing was higher in the intervention arm (98% versus 92%, difference +6%, 95%CI +2% to +10%, p=0.004). Yield of HIV testing was 2.1% in the intervention arm and 0.6% in the control arm (p=0.22).

**Conclusion:** A norming intervention significantly improved uptake of HIV testing among household contacts of persons with TB.

**Funding/Support:** This work was supported by the Center for Interdisciplinary Research on AIDS (P30MH062294) and the Fogarty International Center of the National Institutes of Health (R21TW011270). The content is solely the responsibility of the authors and does not necessarily represent the official views of the NIH or other sponsors.

## Introduction

HIV status awareness and linkage to care are critical for ending the HIV epidemic and preventing tuberculosis (TB).^1^ This is especially critical for close contacts of persons with TB, for whom HIV greatly increases the risk of prevalent TB^2^ and incident TB and death.^3^ TB remains the leading cause of death among people living with HIV (PLHIV).^4,5^ Yet layered or intersecting stigma for HIV and TB may create substantial barriers to seeking HIV testing at clinics for close contacts of persons with TB.

Nesting home-based HIV testing into TB contact investigation provides high-risk and hard-to-reach populations an opportunity to test for HIV. Home-based HIV testing can reach individuals outside the health system, eliminate the costs of attending clinics for testing and provide testing in a familiar, private environment. Moreover, home-based HIV testing can be delivered by specialized lay health workers trained to provide HIV counseling, supervised testing, and linkage to clinic-based care.^6^ However, up to half of household contacts decline HIV test offers during contact investigation, with contacts citing concerns that others in the household may think less of them if they test as a primary reason they hesitate to test, even when counseling and testing is free, accessible, and confidential.^7^ The perception that testing for HIV is uncommon in one’s social network may increase perceived stigma and decrease willingness to test for HIV.

Growing evidence suggests that misperceived norms influence HIV-related health behaviors, including the willingness to test for HIV.^8^ A ‘norming’ intervention to facilitate re-evaluation of expectations related to HIV testing may increase uptake of testing by exposing individuals to the attitudes, values and behaviors of household members who support HIV testing.^9^ We evaluated a brief, community health worker-delivered norming strategy to increase acceptance of HIV testing during TB household contact investigation compared to standard strategies.

## Methods

### Study design and setting

We carried out a household-randomized, controlled trial to evaluate the effect of a norming strategy among household contacts of persons with pulmonary TB in Kampala, Uganda (ClinicalTrials.gov #NCT05124665).

### Setting

The prevalence of HIV in Uganda is estimated to be 5.4% and TB incidence to be 200 per 100,000.^10^ Over 25% of PLHIV in Uganda are unaware of their HIV status.^11^ In previous studies, only 53-61% of household contacts of persons with TB accepted offers of free, optional home-based HIV testing.^6,12,13^ For this study, we chose TB clinics at three public Kampala Capital City Authority (KCCA) health facilities in Kampala, Uganda based on each having a high volume of persons with TB, including a substantial proportion of persons also living with HIV. All TB services, including home-based TB contact investigation, are provided free through a partnership with the Uganda Ministry of Health.

### Recruitment and Randomization

Community health workers (CHW) enrolled index persons with TB who were eligible for household contact investigation, reported two or more household contacts aged 15 years or older, and were willing to participate in the study. Households were then randomized to standard-of-care or intervention services using variable block randomization, with block sizes of 2, 4 or 8 to mask the end of a block. Separate teams of CHWs visited households to deliver intervention or standard-of-care services.

### Procedures

CHWs visited homes of index persons with TB in pairs to screen contacts for TB symptoms and offer free, optional, oral HIV testing. Households were eligible for study inclusion if they included at least two contacts ≥15 years who self-reported not to be living with HIV. Regardless of study inclusion or randomization, all contacts were offered free TB contact investigation services and free, optional HIV testing. CHWs collected data on contact demographics; TB symptoms; self-reported HIV status; responses to a validated HIV-TB stigma scale; and HIV testing eligibility, decision, and test outcomes, using a customized, encrypted, electronic data collection system (CommCare, Dimagi, Boston, Massachusetts, USA) on password-protected Android tablets.

Trained CHWs with more than 5 years of experience offering integrated TB and HIV services in the home setting in accordance with Uganda National TB and Leprosy Programme (NTLP) guidelines.^14^ Eligible contacts were individually taken aside to a private place, given HIV counseling, and offered a free, saliva-based HIV test (OraQuick, OraSure Technologies, Bethlehem, Pennsylvania, USA). The order in which eligible contacts were offered the HIV test was left to the CHW’s discretion.

### Intervention

The intervention consisted of a social-behavioral norming strategy to address misperceived norms related to HIV testing. It comprised five components, each designed to influence household dynamics to promote acceptance of HIV counseling and testing. The components were (1) guided selection of a first tester likely to accept testing; (2) use of a prosocial script; (3) opt-out framing of the test offer; (4) optional sharing of decisions to test and (5) masking decisions not to test. These components were delivered by a CHW during a single home visit, guided by decision support prompts and scripts integrated into the electronic case record application. Intervention components are summarized in **Figure 1** and the rationale for each is described in the trial protocol.

**Figure 1:**
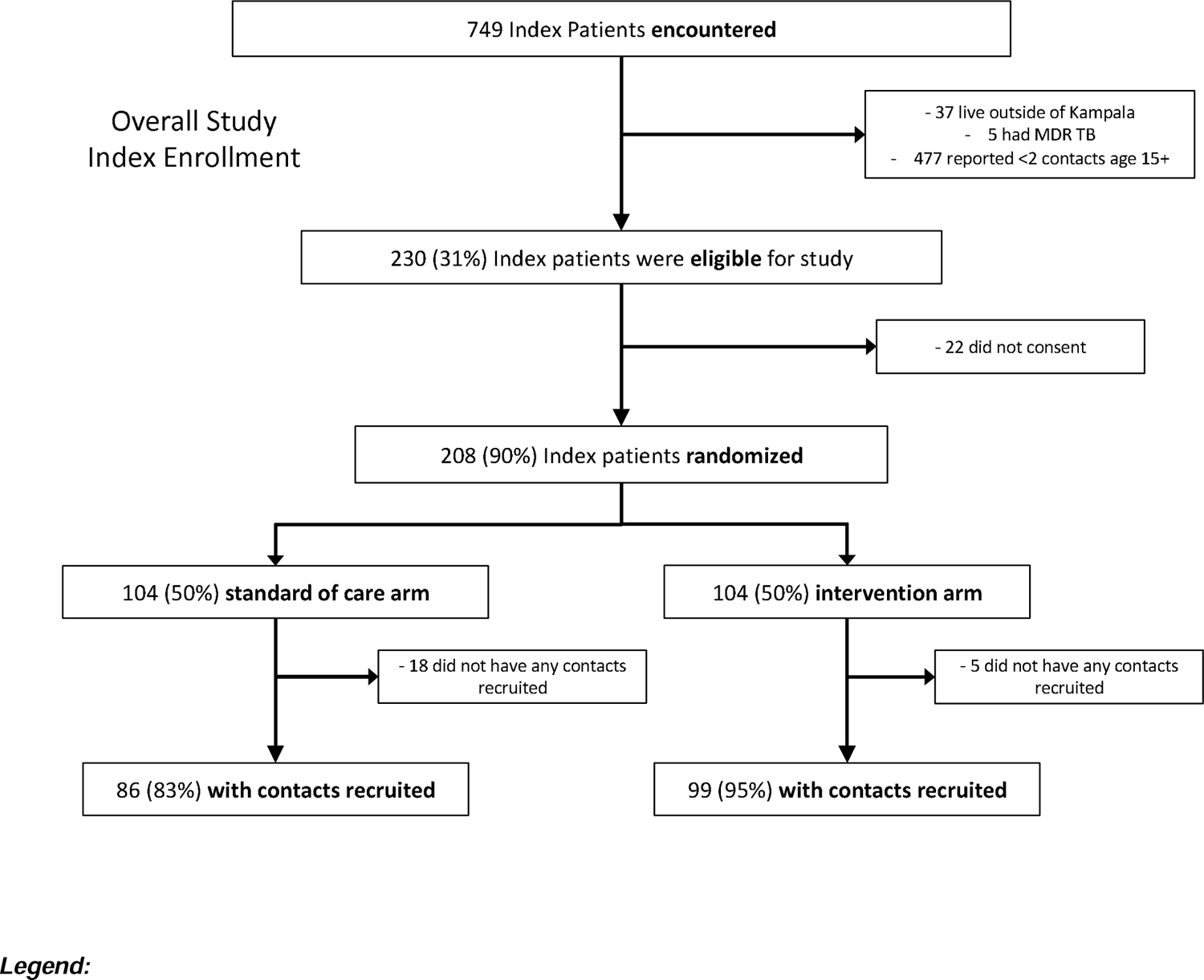
CONSORT diagram

**Figure 1:**
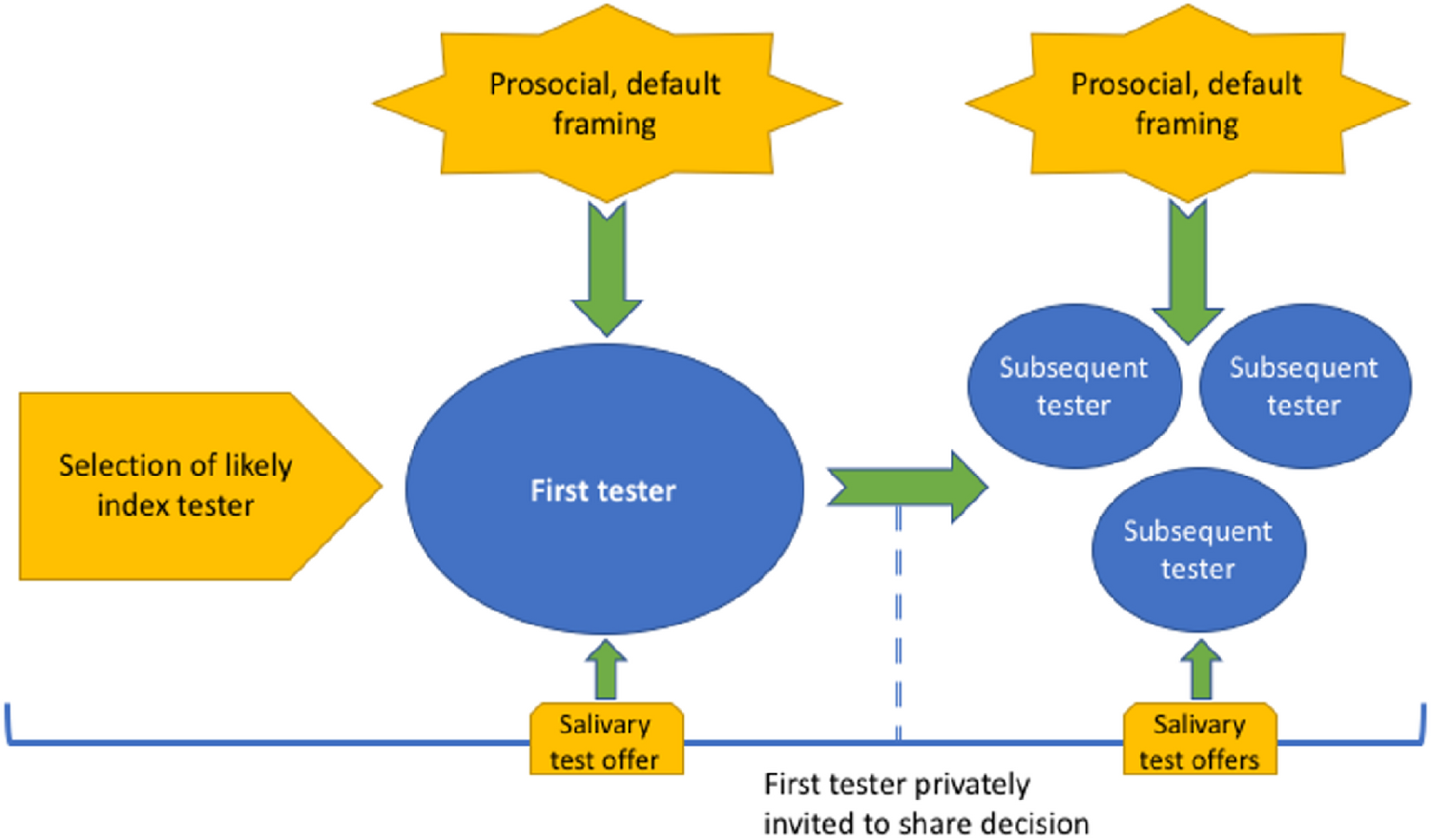
Diagram of intervention components

In households assigned to the intervention, the CHW offered HIV testing first to the oldest female present. In introducing the HIV test offer, the CHW explained that deciding to test for HIV might empower others in the participant’s household to test as well. If the participant decided to complete an HIV test, they were offered the free, saliva-based HIV test. After receiving their results, they were also invited to share their decision to test with the household. If the participant decided not to complete an HIV test, they were counseled about other options for accessing HIV testing and asked to complete a short survey comparable in duration to HIV testing. Regardless of testing decision, all participants spent approximately equal periods of time with the CHW to prevent inadvertent disclosure of testing choice. After offering HIV counseling and testing to the first household contact, the CHW team repeated the offers to each eligible member of the household.

### Measures

The primary outcome was the proportion of contacts accepting HIV testing, defined as the number of eligible contacts who underwent HIV testing divided by the total number of contacts offered testing. CHWs collected demographic and clinical information, including age, sex, education, and TB symptoms, from participants on electronic case record forms. They assessed perceived stigma related to HIV and TB before HIV test offers using Van Rie scales adapted for Uganda before HIV test offers with possible scores ranging from 0 to 18. Index persons with TB underwent HIV testing by a clinic-based CHW prior to the home visit.

### Sample size calculations

Based on a previous trial of household contact investigation for TB, we assumed an average of two household contacts would be eligible for HIV testing per household, the intraclass correlation coefficient (ICC) within households would be 0.59, the proportion consenting to testing in the control group would be 0.85, and proportion consenting to testing in the intervention group would be 0.98. With these assumptions, we calculated that 152 households would achieve 90% power to detect an effect of +0.13Dat α=0.05. We inflated the target sample size for enrollment by 30% to account for losses to follow-up before home visits, which is common in the setting.^15^

### Statistical analysis

We produced descriptive statistics for the total population and conducted bivariate tests by study arm to assess balance on key variables: age, sex, index HIV status, and reported TB symptoms. The primary outcome, the proportion of contacts accepting HIV testing, was analyzed using an intention-to-treat approach and logistic regression. We fit a mixed-effects model to assess and account for clustering by household. We assessed the ICC and fit a multivariable logistic regression model. We assessed HIV testing yield as a proportion of all contacts tested and compared yield across arms.

### Ethics and role of the funding source

This study was approved by the Yale Human Investigation Committee (#2000024852), the Makerere University School of Public Health Institutional Review Board (#661), and the Uganda National Council on Science and Technology (#HS2567). All participants provided verbal informed consent. The funding sources played no role in the design, conduct, or reporting of this study.

## Results

### Population

Three clinic-based CHWs screened 749 consecutive index persons with TB for eligibility from November 2021 to July 2022 (Figure 1). CHWs enrolled 208 consenting index persons with TB for household visits; 104 households were randomized to the intervention and 104 to the standard-of-care arm.

Two field-based CHWs enrolled 328 contacts from 99 index households in the intervention arm and 224 contacts from 86 index households in the standard-of-care arm (**Figure 1**). In the intervention arm, 285 (87%) contacts met eligibility criteria. In the control arm, 187 (84%) contacts met eligibility criteria.

Intervention and control arms were balanced by age, gender, education level, and years spent living in current home (**Table 1**). 94 (33.0%) contacts in the intervention arm and 29 (15.5%) contacts in the control arm were living with an HIV-positive index person with TB (p<0.001).). Baseline perceived stigma for HIV within the household measured before HIV test offers was higher in the intervention arm than in the control arm (mean stigma score 9.5 versus 7.7, p=0.003).

**Table 1:** Characteristics of household contacts.

|  | **Standard-**<br>**of-care** | **Intervention** |
| --- | --- | --- |
|  | **N=187** | **N=285** |
| **Age<sup>+</sup>** | 31 (14) | 30 (13) |
| **Male** | 65 (34.8) | 98 (34.4) |
| **Index living with HIV** | 29 (15.5) | 94 (33.0) |
| **Education** |  |  |
| No formal education | 7 (3.7) | 13 (4.6) |
| 1-4 | 15 (8.0) | 22 (7.7) |
| 1-7 | 50 (26.7) | 66 (23.2) |
| O-Level | 66 (35.3) | 104 (36.5) |
| A-Level | 18 (9.6) | 38 (13.3) |
| Vocational | 5 (2.7) | 5 (1.8) |
| Tertiary/university | 24 (12.8) | 31 (10.9) |
| Post-graduate | 2 (1.1) | 6 (2.1) |
| **Years living in home** | 8.7 (9.7) | 9.9 (10.8) |
| **Baseline stigma score** | 7.7 (6.5) | 9.5 (6.2) |
| **Symptoms of TB** |  |  |
| Coughing | 35 (18.7) | 46 (16.1) |
| Fever | 24 (12.8) | 13 (4.6) |
| Weight loss | 18 (9.6) | 16 (5.6) |
| Night sweats | 15 (8.0) | 14 (4.9) |
| **Tested for HIV<sup>+</sup>** | 172 (92.0) | 280 (98.3) |
| **Tested positive for HIV** | 1 (0.6) | 6 (2.1) |

<sup>+</sup> N (%) unless otherwise specified.
+Mean (SD)

### Acceptance of HIV testing

Four hundred fifty-two contacts accepted and completed HIV testing during the home visit. The intra-cluster correlation coefficient by household was very low at 0.016. Acceptance and completion of HIV testing were higher in the intervention arm (98% versus 92%, difference +6%, 95%CI +2% to +10%, p=0.006) (**Table 2**). The odds of a contact accepting HIV testing in the intervention arm were 4.6 times the odds of a contact completing a test in the control arm (95% CI 1.6-12.9, p=0.004). The adjusted risk difference between arms was 6%. A multivariable model adjusting for index person HIV status resulted in similar effect estimates and confidence intervals (**Table 2**).

**Table 2:** Completion of HIV testing.

|  | **Risk difference** | **95% confidence interval** | **p value** |
| --- | --- | --- | --- |
| **Primary model\*** |  |  |  |
| Intervention<sup>†</sup> | +5.7% | 1.7% - 9.9% | 0.006 |
| **Multivariable model with index HIV status<sup>‡</sup>** |  |  |  |
| Intervention | +5.5% | 1.4% - 9.6% | 0.008 |

\*Results are from a logistic regression model. A mixed effects model accounting for clustering by household showed a very low intra-cluster correlation coefficient (ICC) of 0.016 and identical parameter estimates and confidence intervals.
<sup>†</sup>Compared to contacts randomized to standard-of-care
<sup>‡</sup>Results are from a multivariable logistic model adjusting for Index Patient HIV status.

### Yield of HIV testing

A total of seven contacts tested positive for HIV during the home visit, including six in the intervention arm and one in the control arm. Yield of HIV testing was 2.1% in the intervention arm and 0.6% in the control arm (p=0.22). All newly found individuals living with HIV were linked to care and initiated on ART. Odds of positive tests among contacts were similar regardless of HIV status of the index person with TB (OR 1.2, 95% CI 0.72-1.9, p=0.53).

## Discussion

Home-based HIV testing can reach key populations, but many household contacts of persons with TB decline HIV tests. We evaluated a simple, brief strategy for improving the uptake of HIV testing among individuals living with an index person with TB by establishing HIV testing as normative. We found that the intervention strategy significantly improved acceptance and completion of HIV testing compared to a standard strategy delivered by experienced CHWs.

In our study, individuals offered HIV testing using the norming strategy were more likely to complete a test than those offered HIV testing using a standard strategy. This finding is consistent with an emerging body of work suggesting that the perception that testing for HIV is normative is associated with increased willingness to test for HIV oneself.^16^ Even in settings where HIV testing is normative, many individuals misperceive HIV testing to be uncommon. In observational studies, the misperception that testing for HIV is rare is associated with reduced willingness to test for HIV. Our study expands on the study of misperceived norms by evaluating a simple strategy to preempt such norms in a randomized controlled trial. Importantly, our study shows that CHWs can intervene on misperceived norms and that doing so is associated with significant increases in uptake of HIV testing.

We also found that the yield of HIV test offers in households offered testing using a norming strategy was 2.1%, while the yield of HIV test offers in households offered testing using a standard strategy was 0.6%. This difference was not statistically significant, and the trial was not powered to detect a difference in yield. However, this finding points to the potential that interventions to correct misperceived norms around HIV testing might disproportionately benefit individuals at higher risk of undetected HIV. One potential mechanism for this improvement may be that the norming intervention empowers harder-to-reach populations to take the opportunity to test. In other words, the individuals who opt to test under the norming strategy who would not have tested under a standard strategy may be less likely to have tested before or be aware of their current status. Future studies should be powered to evaluate whether the norming strategy improves the yield of HIV testing and linkage to care.

### Limitations

This study had some limitations. First, our study was powered only to detect an effect on completion of HIV testing, not on yield of HIV testing, linkage to care, or initiation of ART. The purpose of this pilot study was to provide a proof-of-concept for an intervention to change uptake of HIV-related services using a norming strategy. Larger, pragmatic trials of norming interventions are warranted. Second, we used variable-block, concealed allocation randomization without stratification by any household or index person characteristics. This approach allowed an imbalance in HIV status among index persons with TB and total enrolled contacts to emerge between study arms. However, adjustment for index HIV status in models demonstrates that this imbalance did not affect the primary outcome. Nonetheless, future studies should stratify by index HIV status and total household size to ensure balance.

This study also has several strengths. To our knowledge, this is the first randomized, controlled trial of an intervention to improve the uptake of HIV testing by groups at high risk of HIV by correcting misperceived norms. Our approach to designing the intervention combined a review of the fast-accumulating social-behavioral research on misperceived norms with best practices for participatory co-design with affected populations. Our approach to evaluating the intervention employed a household-randomized, controlled design with block randomization and separate CHW teams from equivalently trained and experienced pools. Finally, we offered both the intervention and usual care groups an oral, saliva-based HIV test that our preliminary research found to be not only acceptable but strongly preferable to participants. Offering the most appealing modality of HIV testing to all participants is a conservative approach to evaluating the effect of the norming intervention because it likely increases the uptake of HIV testing for the usual care comparison group.

## Conclusion

Norming strategies are simple, inexpensive, and ideal for delivery by CHWs. We evaluated a simple, brief strategy for improving the uptake of HIV testing among individuals living with an index person with TB. We found that the intervention strategy increased acceptance and completion of HIV testing compared to a standard strategy delivered by experienced CHWs. This finding is consistent with an emerging body of work suggesting that the perception that testing for HIV is normative is associated with increased willingness to test for HIV oneself. Larger randomized controlled trials are needed to evaluate the effect and cost-effectiveness of norming interventions on HIV testing yield, linkage to care, and ART initiation.

## Data Availability

All data produced in the present study are available upon reasonable request to the authors

## Author Contributions

Study concept and design: MAH, LD

**Collection of data:** JG, JN

Analysis and interpretation of data: **MAH, AJG**

**Drafting of the manuscript:** MAH

**Review of the manuscript**: JG, AJG, JN, AK, LD

**Statistical analysis:** MAH, AJG

Obtained funding: **LD**

**Conflict of interest disclosures:** No disclosures were reported.

**Funding/Support:** This work was supported by the National Institutes of Health under Award R21TW011270 (JLD). The content is solely the responsibility of the authors and does not necessarily represent the official views of the National Institutes of Health.

**Role of the sponsors:** The funding organizations had no role in the design and conduct of the study; in the collection, analysis, and interpretation of the data; or in the preparation or approval of the manuscript.

**Data sharing:** De-identified minimally reproducible data and code are available by request.

**Disclaimer:** This manuscript does not necessarily represent the view of the U.S. Government.

## Notes

### Competing Interest Statement

The authors have declared no competing interest.

### Clinical Trial

NCT05124665

### Author Declarations

Ethics committee/IRB of Yale University gave ethical approval for this work.

## References

1. HIV/AIDS JUNPo. Fast-track: ending the AIDS epidemic by 2030. 2014.

2. Velen K, Shingde RV, Ho J, Fox GJ. The effectiveness of contact investigation among contacts of tuberculosis patients: a systematic review and meta-analysis. Eur Respir J. Dec 2021;58(6)doi:10.1183/13993003.00266-2021

3. Hopewell PC. Impact of human immunodeficiency virus infection on the epidemiology, clinical features, management, and control of tuberculosis. Clin Infect Dis. Sep 1992;15(3):540–7. doi:10.1093/clind/15.3.540

4. World Health Organization. Global Tuberculosis Control: WHO Report 2022. 2022.

5. UNAIDS. Global AIDS Update 2022. 2022.

6. Ochom E, Meyer AJ, Armstrong-Hough M, et al. Integrating home HIV counselling and testing into household TB contact investigation: a mixed-methods study. Public Health Action. // 2018;8(2):72–78. doi:10.5588/pha.18.0014

7. Armstrong-Hough M, Ggita J, Ayakaka I, et al. Brief Report: “Give Me Some Time”: Facilitators of and Barriers to Uptake of Home-Based HIV Testing During Household Contact Investigation for Tuberculosis in Kampala, Uganda. J Acquir Immune Defic Syndr. Apr 1 2018;77(4):400–404. doi:10.1097/qai.0000000000001617

8. Perkins JM, Kakuhikire B, Baguma C, et al. Perceptions About Local ART Adherence Norms and Personal Adherence Behavior Among Adults Living with HIV in Rural Uganda. AIDS Behav. Jan 16 2022;doi:10.1007/s10461-021-03540-1

9. Armstrong-Hough M, Ggita J, Gupta AJ, et al. Assessing a norming intervention to promote acceptance of HIV testing and reduce stigma during household tuberculosis contact investigation: protocol for a cluster-randomised trial. BMJ Open. May 25 2022;12(5):e061508. doi:10.1136/bmjopen-2022-061508

10. UNAIDS. Country Factsheet: Uganda. 2022.

11. WHO. Technical report: Assessment of HIV testing services and antiretroviral therapy service disruptions in the context of COVID-19: Lessons learned and way forward in sub-Saharan Africa. 2021. https://www.who.int/publications/i/item/9789240039599

12. Davis JL, Turimumahoro P, Meyer AJ, et al. Home-based tuberculosis contact investigation in Uganda: a household randomised trial. ERJ Open Res. Jul 2019;5(3)doi:10.1183/23120541.00112-2019

13. Armstrong-Hough M, Ggita J, Turimumahoro P, et al. ‘Something so hard’: a mixed-methods study of home sputum collection for tuberculosis contact investigation in Uganda. Int J Tuberc Lung Dis. Oct 1 2018;22(10):1152–1159. doi:10.5588/ijtld.18.0129

14. Health UMo. Manual for Management and Control of Tuberculosis and Leprosy. 2017.

15. Armstrong-Hough M, Turimumahoro P, Meyer AJ, et al. Drop-out from the tuberculosis contact investigation cascade in a routine public health setting in urban Uganda: A prospective, multi-center study. PLoS One. 2017;12(11):e0187145. doi:10.1371/journal.pone.0187145

16. Perkins JM, Nyakato VN, Kakuhikire B, et al. Actual Versus Perceived HIV Testing Norms, and Personal HIV Testing Uptake: A Cross-Sectional, Population-Based Study in Rural Uganda. AIDS Behav. Feb 2018;22(2):616–628. doi:10.1007/s10461-017-1691-z

